# Percentage HScore confirms low incidence of secondary haemophagocytic lymphohistiocytosis in hospitalised COVID-19 patients

**DOI:** 10.1101/2020.10.19.20214015

**Authors:** Michael R Ardern-Jones, Matt Stammers, Hang T.T. Phan, Florina Borca, Anastasia Koutalopoulou, Ying Teo, James Batchelor, Trevor Smith, Andrew S Duncombe

## Abstract

**Objective:** It has been assumed that a significant proportion of COVID-19 patients show evidence of hyperinflammation of which secondary haemophagocytic lymphohistiocytosis (sHLH) is the most severe manifestation. To facilitate diagnosis of sHLH the HScore has been developed and validated. We set out to examine the prevalence of sHLH-like hyperinflammation in COVID-19.

**Methods:** We retrospectively examined HScore parameters in 626 COVID-19 cases admitted to our institute of which 567 were suitable for analysis and compared these to a cohort of confirmed infection associated sHLH cases. To account for missing data, we calculated the maximum possible HScore of the recorded parameters (%HScore).

**Results:** Early measurement of HScore parameters (day -1 to 4 from diagnosis) strongly predicted the %HScore over the course of the admission (p <0.0001). The retrospective cohort of sHLH showed significantly higher %HScores as compared to COVID-19 (median 73.47 vs 18.13 respectively, p <0.0001). The overall prevalence of individuals with an 80% probability of sHLH in our COVID-19 cohort was 1.59% on admission and only rose to 4.05% during the whole disease course. In the small cohort with scores suggestive of sHLH, there was no excess mortality compared with the whole cohort. %HScores were higher in younger patients (p<0.0001) and did not reliably predict outcome at any cut-off value (AUROC 0.533, p=0.211; OR 0.99).

**Conclusion:** Surprisingly, these findings show that sHLH-type hyperinflammation is not prevalent in COVID-19, and %HScores do not predict outcome. Therefore, new algorithms are required to optimise case selection for clinical trials of targeted anti-inflammatory interventions.

## Introduction

Mortality from SARS-CoV-2 infection causing COVID-19 in hospitalised patients in the United Kingdom has been reported to be 25.7% (1). The principal cause of death due to COVID-19 is respiratory failure due to acute respiratory distress syndrome (2). Early reports have suggested that a subgroup of individuals suffer a hyperinflammatory state with high mortality which is associated with high levels of IL-6 and CRP (3). Hyperinflammation has been previously described secondary to acute infection and termed cytokine release syndrome / cytokine storm (CRS / CS), macrophage activation syndrome (MAS), macrophage-cytokine self-amplifying loop (MCSAL) and secondary haemophagocytic lymphohistiocytosis (sHLH). Viral infections are the commonest cause of sHLH (4), and symptoms of hyperinflammation resemble those of general sepsis, therefore hyperinflammation has generally been under-recognised at an early stage leading to high mortality (5). Recent randomised controlled trial data has shown that the anti-inflammatory agent dexamethasone can reduce mortality in severe COVID19 in an unselected COVID-19 cohort(1). Whilst impressive, these results suggest that if the anti-inflammatory interventions could be targeted early to individuals with hyperinflammation, even greater benefit in mortality may be seen, and this approach may additionally reduce the morbidity of COVID-19 by preventing escalation to high dependency and intensive care. Therefore, it is likely that strategies to identify hyperinflammation and targeted intervention will offer the most effective approach to the management of hyperinflammation in COVID-19. Indeed, as well as anti-IL-6 (6) other cytokines released in hyperinflammation for which existing biologic therapies are available are also potential targets for intervention including TNF-α (Infliximab), IL-1 (Anakinra), and JAK-inhibitors (e.g. Ruxolitinib).

To facilitate diagnosis of hyperinflammation, the ‘HScore’ (7, 8) has been developed in the context of sHLH because of evidence that early recognition and intervention is beneficial (Supplementary Table 1)(8). Some authors have recommended using the HScore in COVID19 to identify hyperinflammation (9). However, others have reported that very few COVID patients fulfil the current HScore criteria (10). This discrepancy may in part be due to missing data for the parameters in the HScore in COVID-19 patients. We have devised a modified HScore expressed as a percentage of the maximum achievable score (%HScore) from the data parameters available for each individual patient in a large dataset of patients admitted to a single institution with a confirmed diagnosis of COVID-19 to further examine its relevance (Supplementary Table 1).

**Table 1.** A table to show the baseline characteristics of the Southampton COVID-19 cohort. IQR, interquartile range; BAME, Black and ethnic minority, BMI, Body mass index.

|  | COVID-19 (n=567) |
| --- | --- |
| Age (Median years, IQR) | 71 (54 - 82) |
| Sex: male (%), female (%) | 58.20, 41.80 |
| BAME (%) | 11.46 |
| BMI (Mean, IQR) | 25.73 (22.49-30.22) |
| Cardiac disease (%) | 71.78 |
| Renal disease (%) | 59.96 |
| Respiratory disease (%) | 41.09 |
| Gastrointestinal disease (%) | 38.10 |
| Diabetes (%) | 26.28 |
| Neurological disease (%) | 51.15 |
| Cancer history (%) | 28.04 |

## Patients and Methods

Following ethical approval (NRES 286016), we recruited all cases of COVID-19 infection that tested positive for SARS-CoV-2 viral RNA in our laboratory and were admitted to University Hospitals Southampton NHS foundation trust between 07/03/2020 and 09/06/2020, n=626. Additionally, we recruited a retrospective cohort of sHLH (infection associated) from the same institution based on confirmed diagnosis recorded as ICD-10 D76.2 (n=16).

Structured and semi-structured data was accrued from the trust integration engine using SQL Developer 4.2 queries and then cleaned/transformed using python 3.7 and associated libraries: *numpy* and *pandas*. Analysis was performed using *matplotlib, seaborn* and *scipy*. Statistical analysis was undertaken using GraphPad, Prism (8.4.3).

The HScore, (Supplementary Table 1), includes 3 clinical parameters (immunosuppression, pyrexia, organomegaly), 5 blood tests (triglyceride, ferritin, transaminase, fibrinogen, cytopenia), and bone marrow aspirate features. Each of these is weighted by variable and a score based on the value/result is summated to provide an overall score from 0 to 337. This value is then utilised to calculate a probability of HLH e.g. HScore of ≤90 probability sHLH <1%, to >99% probability with an HScore of ≥250. We calculated the HScore based on parameters available retrospectively. As expected from the infective precautions taken on COVID-19 patients, little data was available on palpable hepatosplenomegaly, bone marrow aspirate histology or immunosuppression and we excluded these three parameters. To account for these missing values we created a modified HScore calculated from the percentage points from the available parameters expressed as a percentage (%HScore, Supplementary Table 1).

The primary outcome utilised in this study was binary: discharge from hospital or death in hospital. Admission date was an unreliable marker of disease onset as some of our cohort contracted COVID-19 after prolonged periods in hospital and therefore the time of initial infection was unclear. Clinical teams arranged testing as symptoms presented and therefore, to facilitate comparison between cases, investigation parameters were normalised to the date of SARS-CoV-2 viral RNA laboratory confirmation and outcome data tabulated from day -1 to day 21.

## Results

The characteristics of the 567 eligible cases (41.8% female) showed a high prevalence of comorbidities in line with the high overall average age (median 71 years; IQR 54-82), (Table 1). In light of the influence of age on mortality, we examined our dataset for the number of recorded HScore parameters (day -1 to 4), as distributed by age (n=621) (Fig 1a). This showed that the individuals where few data points were available, were more likely to be older (p=0.0025). To address this source of potential bias, we removed individuals with fewer than three data points from further analysis. Subsequent analysis of the distribution of data points in the reduced cohort (n=567) confirmed no association between the number of data parameters and age (p=0.094). In order to determine the role of %HScore for early identification of hyperinflammation, we therefore restricted further analysis to day -1 to 4. We then determined that %HScore measured in the first 5 days of illness (day -1 to 4 after laboratory virus confirmation) was a very strong predictor of the %HScore during the whole admission (r=0.8499, p<0.0001, Fig 1b).

**Figure 1.**
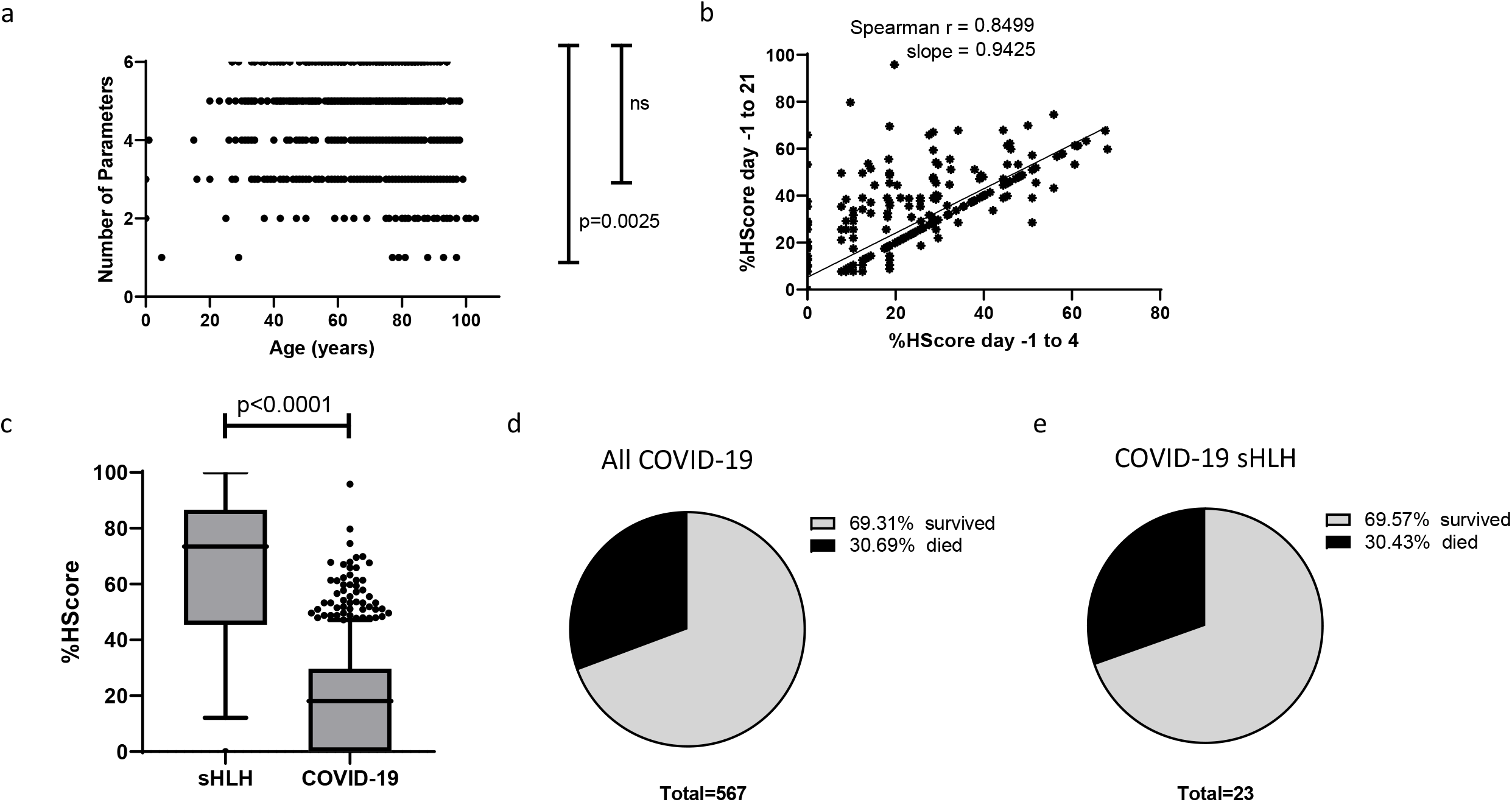
sHLH shows a higher %HScore than COVID-19, but %HScore in COVID-19 shows no correlation with mortality. a. Number of HScore parameters available for analysis in the dataset per age of the patient. One-way Anova. n= 621 b. %HScore as measured from data points recorded at virus diagnosis timepoint day -1 to 4, versus day -1 to 21. Spearman’s correlation coefficient (r) presented. n = 567 c. Plot of %HScores from a retrospective cohort of secondary haemophagocytic lymphohistiocytosis (sHLH) versus COVID-19. Error bars represent 10-90% confidence. Mann Whitney test presented. d. d. Pie chart showing proportion of patients with COVID-19 who survive or die. e. Pie chart showing proportion of patients with a %HScore ≥56.7 (80 % probability of sHLH) who survive or die

To determine the prevalence of HLH-like disease in the COVID-19 cohort we examined the %HScore in our cohort versus a retrospective cohort of sHLH cases also from our institution. Comparing sHLH cases with COVID-19, we found that %HScores were significantly higher (median 73.47 vs 18.13 respectively, p <0.0001, Fig 1c). An HScore which predicts an 80% probability of sHLH is reported to be 191/337 (7) which is equivalent to a %HScore of 56.7%. These criteria were met by 75% of sHLH cases but only 1.59% (9 of 567) COVID-19 cases. If %HScore was calculated from ‘worst’ values at any time day -1 to 21, the proportion of COVID-19 cases meeting the sHLH threshold was only marginally higher at 4.05% (23 of 567). For those individuals with a %HScore above the sHLH threshold, there was no increase in mortality as compared to the whole cohort mortality of 30.69% (p >0.05) (Fig 1d, e).

Overall mortality was strongly predicted by patient age (p <0.0001; median age survivors 64 years IQR 49-76; died 81 years, IQR 73 – 87; Fig. 2a). At a threshold of 75 years of age, the increased risk of mortality was significant (OR 7.295, 4.89 – 10.8, p<0.0001). However, age conferred a strong negative correlation on %HScore (Spearman r = -0.305, -0.38 to -0.226, p < 0.0001; Fig 2b), across the cohort. Strikingly, the median %HScore was significantly lower (p < 0.0001) in the older age group: >75 years median %HScore 7.724 (0.0 to 18.16) vs ≤ 75 years median %HScore 18.31 (7.72 to 28.57) (Fig 2c). Receiver operator characteristics (ROC) over the whole cohort suggest that at any threshold, %HScore is not useful as a predictor of mortality in COVID-19 (AUROC 0.533, p=0.211; OR 0.99, 0.98 to 1.00) (Fig 2d). However, because of the very strong association between age and mortality, it is important to stratify for age to examine the effect of %HScore on mortality. This showed that the negative correlation between age and %HScore was highly significant in both those who survived (r = -0.307, -0.441 to -0.164, p <0.0001) and those who died (r = -0.309, -0.441 to - 0.164, p<0.0001) and that there was no difference in %HScore between those who died and survived (p = 0.3125) (Fig 2e).

**Figure 2.**
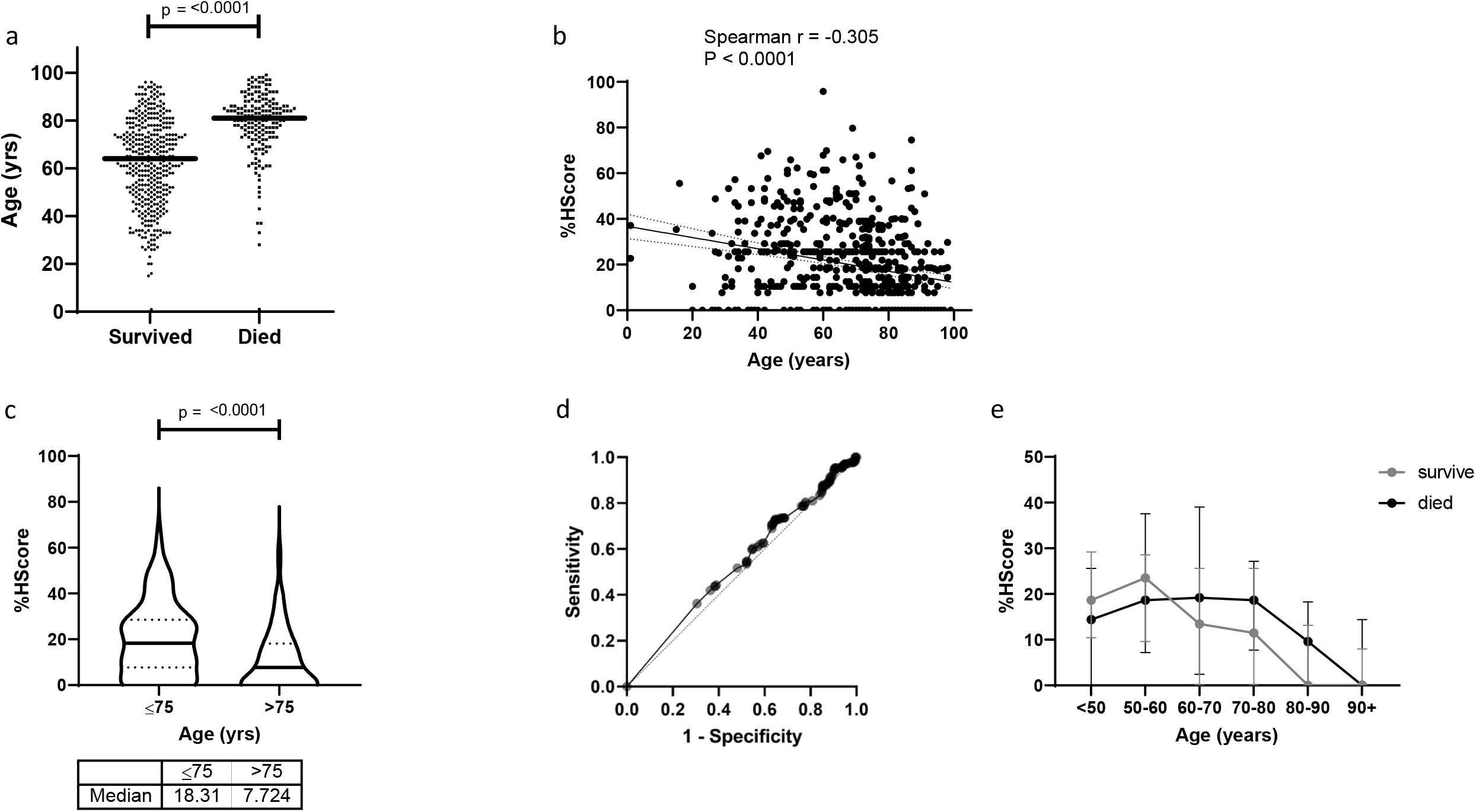
Age and risk of mortality in COVID-19. a. Scatter column plot of COVID-19 cases showing age (years) of the subgroups who survived versus those who died (n=567). Horizontal bars represent median value. Mann Whitney statistic presented. b. Scatterplot of COVID-19 cases showing age versus %HScore. Linear regression (dark line) with 95% confidence limits (dotted lines), with Spearman’s correlation coefficient (r) and p statistic presented. c. Violin plot of %HScore in those ≤ 75 versus >75 years (n = 567). Horizontal bars represent median value, interquartile range dotted. Mann Whitney statistic presented. d. Receiver operator characteristics of prediction of mortality by %HScore e. %HScore in cases who died (black dots) versus survived (grey dots) by age stratification. Error bars represent 25% - 75% confidence interval.

## Discussion

The limitations of this work include the retrospective design and heterogeneity across the cohort of the recorded number of HScore parameters for assessment. However, the design allowed us to recruit all cases of viral RNA confirmed COVID-19 cases and we report here one of the largest datasets of COVID-19 to date which exceeds the 312 sHLH cases in the original series identifying the HScore (7) and the 40 cases where HScore was applied to intensive care patients (11). To address missing data, we utilised a mathematical programmed approach to facilitate rigorous data collection from centralised hospital electronic records and utilised cross-checking and cross-validation to optimise data cleaning, thus avoiding collection errors, while minimising missing data. Furthermore, to identify the subgroup with sHLH in COVID-19 we undertook a stringent approach to the analysis and did not impute any missing values and instead designed a modified HScore, %Hscore.

In this report, we demonstrate that sHLH is rare in hospitalised cases of COVID-19 similar to the reports of low incidence in intensive care settings (9, 11). Indeed, we estimate that sHLH arises in 1.59% of hospitalised COVID-19 cases early in the course of the illness, and only rising to 4.7% over the whole admission. Surprisingly, mortality in the cohort of COVID-19 cases meeting 80% probability of sHLH showed no excess mortality as compared to the whole cohort (30.43% vs 30.69%). It is notable that the index cohort of sHLH cases used to define the HScore had a median age of 51 years (IQR 36–64) (7), as compared to our COVID-19 patients whose median age was 71 years (IQR 54 -82). In addition, we identified that younger patients have significantly higher %HScores (p<0.0001) and additionally show that when stratified for age, there was no difference in %HScore. Why %HScore (and HScore parameters) decline with age in the context of COVID-19, is not clear and may predominantly reflect immunosenescence. In part this may be explained by responses to COVID, generally acting in an opposite direction to HLH. For example, while pancytopaenia would produce a higher %HScore, it seems that responses to the virus in older individuals are more likely to show increases in circulating white blood cells and platelets, which would clearly drive the %HScore down. Therefore, the association between reduced %HScore and age, as well as the relatively low mortality of sHLH in COVID-19, suggests that waning immunity with age may actually be protective against sHLH-type responses in COVID-19.

IL-6 has been recognised as a key cytokine in COVID-19 (12) and also an important regulator of liver inflammation (13). Fever is regulated by IL-6 and other cytokines including interleukin (IL)-1 and tumour necrosis factor (TNF)-α(14). Dysregulation of these cytokines in older age(15) may further contribute to the lack of sensitivity of measures of the acute phase response in %HScore including temperature and ferritin.

Although, it is possible that high %HScores in COVID-19 do reflect dysregulated immunity, the absolute difference between those who die and survive is small, suggesting that the individual with a high %HScore may lie at or close to a tipping point between harm and benefit from innate inflammation. Therefore, it remains unclear what the effect of broadly applied anti-inflammatory therapies will have on older individuals in particular and a careful balance needs to be struck when designing clinical trials of anti-inflammatory therapies to determine where an individual lies on the risk spectrum of an excessive inflammatory response versus an impaired anti-viral response. Improved endotyping of COVID-19 cases by classification of validated biochemical and molecular phenotypes to identify the subgroup who will benefit from anti-hyperinflammation strategies is critical and these should be used to stratify COVID-19 patients in the next phase of clinical trials.

In summary, we present data which shows that the HScore parameters as measured by a modified scale (%HScore) early in the disease course of hospitalised COVID-19 patients, are of little value for risk stratification with regards to mortality. Whilst those who suffer a worse outcome may show signs of increased inflammation, anti-inflammatory treatments are likely to be detrimental in those without hyperinflammation and therefore we believe that a more sensitive analysis of hyperinflammation biomarkers is urgently required.

## Data Availability

Data sharing will be considered upon request

## Notes

Authors Declaration: No financial support or other benefits from commercial sources for the work reported on in the manuscript, or any other financial interests are declared. No potential conflict of interest or the appearance of a conflict of interest with regard to the work declared.

### Competing Interest Statement

The authors have declared no competing interest.

### Funding Statement

None

### Author Declarations

NRES 286016 This is the reference number for the approval obtained from UK Research Ethics Service (RES) of the national Health Research Authority https://www.hra.nhs.uk/about-us/committees-and-services/res-and-recs/research-ethics-service/

