## Supplementary Table 1 for "Percentage HScore confirms low incidence of secondary haemophagocytic lymphohistiocytosis in hospitalised COVID-19 patients"

Article Type: Brief Report	Running head: COVID HLH

Keywords: Hyperinflammation, SARS-CoV-2, HScore, %HScore, secondary haemophagocytic lymphohistiocytosis, COVID-19

Supplementary Table: 1

Authors Declaration:

No financial support or other benefits from commercial sources for the work reported on in the manuscript, or any other financial interests are declared. No potential conflict of interest or the appearance of a conflict of interest with regard to the work declared.

| **Parameter** | **HScore points (criteria)** | **%HScore points (criteria)**  **(Minimum variables ≥3)** |
| --- | --- | --- |
| Temperature (°C) | 0 (<38.4), 33 (38.4–39.4), or 49 (>39.4) | 0 (<38.4), 33 (38.4–39.4), or 49 (>39.4) |
| No. of cytopenias* | 0 (1 lineage), 24 (2 lineages), or 34 (3 lineages) | 0 (1 lineage), 24 (2 lineages), or 34 (3 lineages) |
| Ferritin (μg/L) | 0 (<2,000), 35 (2,000–6,000), or 50 (>6,000) | 0 (<2,000), 35 (2,000–6,000), or 50 (>6,000) |
| Triglyceride (mmol/L) | 0 (<1.5), 44 (1.5–4), or 64 (>4) | 0 (<1.5), 44 (1.5–4), or 64 (>4) |
| Fibrinogen (g/L) | 0 (>2.5) or 30 (≤2.5) | 0 (>2.5) or 30 (≤2.5) |
| AST/ALT (IU/L) | 0 (<30) or 19 (≥30) | 0 (<30) or 19 (≥30) |
| Hemophagocytosis ** | 0 (no) or 35 (yes) | - |
| Immunosuppression | 0 (no) or 18 (yes) | - |
| Hepatomegaly / Spelnomegaly | 0 (none), 23 (either), or 38 (both) | - |
| **Score** | **Sum of points above (maximum 337)** | **Sum of points above / maximum score (maximum 100%)** |

Supplementary Table 1. HScore and %HScore parameters.

Table to show the differences in between HScore (1) and %HScore.

*haemoglobin ≤92 g/L and/or WBC ≤5 x 10^9^/L and/or platelets ≤110 x 10^9^/L

**features on bone marrow aspirate

AST, Aspartate Transaminase; ALT, Alanine aminotransferase; IU, International Units

1.	Fardet L, Galicier L, Lambotte O, Marzac C, Aumont C, Chahwan D, et al. Development and validation of the HScore, a score for the diagnosis of reactive hemophagocytic syndrome. Arthritis & Rheumatology. 2014;66(9):2613-20.
